# The Long-Term Implications of Childhood Health on Adult Health at age 51

**DOI:** 10.1101/2025.05.14.25327216

**Authors:** Jamie O’Halloran

**Author notes:** Correspondence to: Dr Jamie O’Halloran, Institute of Public Policy Research, 8 Storey’s Gate, Westminster, London.

## Abstract

**Objective:** To examine the association between childhood health and adult health outcomes.

**Methods:** We used data from the 1970 British Cohort Study, which follows participants from age 10 to age 51. Childhood health at age 10 was measured using the Rutter Scale for emotional and behavioural difficulties and self-reported physical health problems that occurred within the last 12 months. We estimated associations using logistic regression models, applying inverse probability weighting to adjust for sample attrition. All models controlled for a wide range of background characteristics reflecting childhood circumstances.

**Results:** We find evidence of associations between health at age 10 and health outcomes at age 51. Severe emotional and behavioural difficulties in childhood were most strongly associated with both the likelihood of experiencing depressive symptoms (relative risk [RR]: 1.85, 95% CI: 1.26–2.56) and with reporting a long-term health condition that affects the amount or type of work (RR: 1.68, 95% CI: 1.19–2.24). Childhood physical health problems were also associated with a higher likelihood of long-term conditions affecting work (RR: 1.38, 95% CI: 1.10–1.68), though we found no statistically significant association with depressive symptoms in midlife.

**Conclusion:** Childhood mental and physical health were associated with adult health outcomes more than four decades later. These findings highlight the potential long-term implications of early-life health for wellbeing in midlife, supporting the case for early intervention and sustained support throughout childhood.

## Introduction

Our early years play a crucial role in shaping our future life outcomes - and our health is no exception. In this analysis, we investigate how mental and physical health issues during childhood correlate to health outcomes into middle age.

This of particular importance in the current context for two key reasons. First, many children in the UK are currently growing up in poor health. Nearly one in four children in England are obese by the time they leave primary school, rising to one in three in the most deprived areas (NHS Digital, 2023a). Alongside this, the UK faces a worsening crisis in children and young people’s mental health. Rates of anxiety, depression, and other common mental health conditions have increased sharply over the past decade, with an estimated one in five children (aged 8 to 15) now experiencing a probable mental disorder (NHS Digital, 2023b).

Second, poor health may be contributing to reduced labour market participation later in life. The UK has comparatively low employment rates among people in midlife (individuals aged 55 to 64) compared to other OECD countries (OECD, 2024). If poor health in childhood is linked to worse economic outcomes in adulthood, this would evidence that if we do not improve the health of our children not only will it have health implications it also may have significant economic ramifications.

This is a well-established area of research, with numerous studies linking childhood health to outcomes such as educational attainment, income, and labour market engagement (e.g. Shlack et al 2021, Chen et al 2024, Case et al 2005, & Goodman et al 2011). Our contribution builds on this evidence base by drawing on new data from the latest wave of the 1970 British Cohort Study. We find that poor mental and physical health in childhood is associated with individuals’ health outcomes more than four decades later.

## Data

We use data from the 1970 British Cohort Study (BCS70), a nationally representative longitudinal study that follows individuals born in a single week in 1970. The cohort has been surveyed approximately every 10 years, with the most recent data capturing outcomes at age 51.

This analysis explores the long-term scarring implications of poor health in childhood, examining how early mental and physical health are associated with later-life health outcomes and their potential implications for wider society. We focus on health at age 10, a key stage in childhood development marking the transition toward adolescence (Sawyer et al 2018) and investigate whether health problems at this age are predictive of outcomes four decades later.

The BCS70 provides rich information on both the childhood experiences of cohort members and the socio-economic circumstances of their families. At age 10, health data were collected through parent reports (typically completed by the mother). The data allow us to control for a wide range of demographic and socio-economic characteristics at childhood when assessing long-term associations.

## Methods

To estimate the association between childhood health at age 10 and health outcomes at age 51, we use logistic regressions. These regressions estimate the increased likelihood of experiencing poor health at age 51 for individuals who had poor physical or mental health in childhood, while controlling for a wide range of potential confounders.

Sample attrition is a common issue for longitudinal surveys – to account for this, we apply inverse probability weighting. Each individual is weighted by the inverse of their estimated probability of responding to the age 51 survey sweep, conditional on having participated at age 10. These probabilities are estimated using a logistic regression model – we use the predictors as identified by Katsoulis et al 2024 that are most strongly associated with non-response in previous BCS70 waves. The full list of covariates used can be found in Appendix Table A1.

To address item non-response in individual characteristics, we use the missing dummy indicator method. For categorical variables, we include a binary indicator for missingness. For continuous variables, we again use the missing indicator method and replace missing values using the gender-specific mean. This approach follows recent studies using the same dataset, such as Foliano et al 2024.

Most of the missingness indicators are statistically insignificant, suggesting that missing data may not be strongly associated with our outcomes. However, while inverse probability weighting and mean imputation with missing-indicator dummies tries to mitigate against potential biases, some residual bias may remain. We discuss this further in the discussion section.

### Measures

We are interested in showing that poor health in childhood can have potential long-term implications on health in adulthood. We investigate two outcomes at age 51 which demonstrate this: self-reported mental health, measured using the Malaise Inventory, and the self-reporting of a long-term physical or mental health condition that affects the amount or type of work that people can do.

The Malaise Inventory is a measure of psychological distress (Rutter 1970), and recent studies have validated its reliability and consistency in capturing symptoms of depression and anxiety in adulthood (Ploubidis et al 2019). A high score (over 8) can be interpreted as experiencing symptoms associated with depression.

Long-term health conditions, whether physical or mental, are important indicators of population health. They are associated with increased healthcare use Marina et al 2020 and can affect labour market outcomes - such as reducing working hours or lowering the likelihood of employment if the condition is severe (Booker et al 2020).

To assess childhood health, we include indicators of both physical and mental health problems, as well as parental health at the time. Childhood mental health is measured using parent-reported data from the Rutter Scale, which captures emotional difficulties, conduct problems, and hyperactivity. We classify the severity of problems using established percentile cut-offs: severe (≥95th percentile), mild/moderate (75th–93rd percentile), and absent (below the 75th percentile) (Rutter et al., 1970).

Physical health in childhood is assessed based on whether the child experienced any health problems in the 12 months prior to the age 10 survey. Following the methodology used by as Parsons et al 2021, we focus on common chronic conditions including eczema, hay fever, wheezing (indicative of asthma), and seizures (‘fits’). Around 20% of children in the cohort experienced at least one of these conditions at age 10.

We also examine the role of parental health in shaping long-term outcomes. Parental mental health is measured using the Malaise Inventory completed by the mother at age 10, and we include indicators of whether either parent had a long-term health problem.

The richness of the BCS70 data allows us to control for a wide range of other factors known to influence health. These include the cohort member’s educational attainment at age 5 and 10, the age the father left education, country of birth, premarital conception, household income at birth, marital status of parents at birth, number of older siblings, socioeconomic status of father, whether the mother smoked during pregnancy.

These covariates allow to control for the circumstance that the child was born into – which is likely correlated to health in childhood whilst not being directly associated with health at age 51.

## Results

For brevity, we report adjusted odds ratios (ORs) for the key variables of interest while controlling for all covariates. The descriptive statistics can be found in Appendix Table A2. Full regression results, including all control variables, are available in Appendix Table A3. The results without covariates are presented in Appendix Table A4.

### Depressive Symptoms at Age 51

Children who experienced emotional and behavioural difficulties at age 10 were more likely to report depressive symptoms at age 51.

Those with severe emotional and behavioural problems had significantly higher odds of experiencing depressive symptoms in midlife. The adjusted odds ratio (aOR) was 2.25 [95% CI: 1.34–3.80], corresponding to a relative risk (RR) of 1.85 [95% CI: 1.26–2.55], indicating an 85% higher risk compared to children with no such difficulties.

Those with moderate symptoms also had elevated odds of experiencing psychological distress in midlife (aOR: 1.32 [95% CI: 0.96–1.78]; RR: 1.25 [95% CI: 0.97–1.56]), though these associations were statistically significant only at the 10% level.

We find no statistically significant association between childhood physical health and depressive symptoms at age 51. However, we find some indication that parental health at age 10 may be associated with depressive symptoms in midlife (aOR: 1.25 [95% CI: 0.97–1.63]; RR: 1.19 [95% CI: 0.98–1.45]), though again, this result is only statistically significant at the 10% level.

### Long-Term Health Conditions at Age 51

Children who experienced emotional and behavioural problems or had physical health conditions at age 10 were significantly more likely to report having a long-term health condition that affects their ability to work at age 51.

Those with severe emotional and behavioural problems had an aOR of 2.01 [95% CI: 1.25–3.22], corresponding to an RR of 1.68 [95% CI: 1.19–2.24], suggesting they were 68% more likely to report a work-limiting long-term health condition compared to those without such difficuties.

Children who had a physical health problem at age 10 also had elevated risk, with an aOR of 1.53 [95% CI: 1.13–2.06] and an RR of 1.38 [95% CI: 1.10–1.68], indicating a 38% higher likelihood of having a work-limiting long-term condition in midlife.

We also find tentative evidence that parental health at age 10 is associated with long-term health outcomes in adulthood (aOR: 1.25 [95% CI: 0.96–1.62]; RR: 1.19 [95% CI: 0.97–1.44]), though this result is only statistically significant at the 10% level.

**Table 1:**
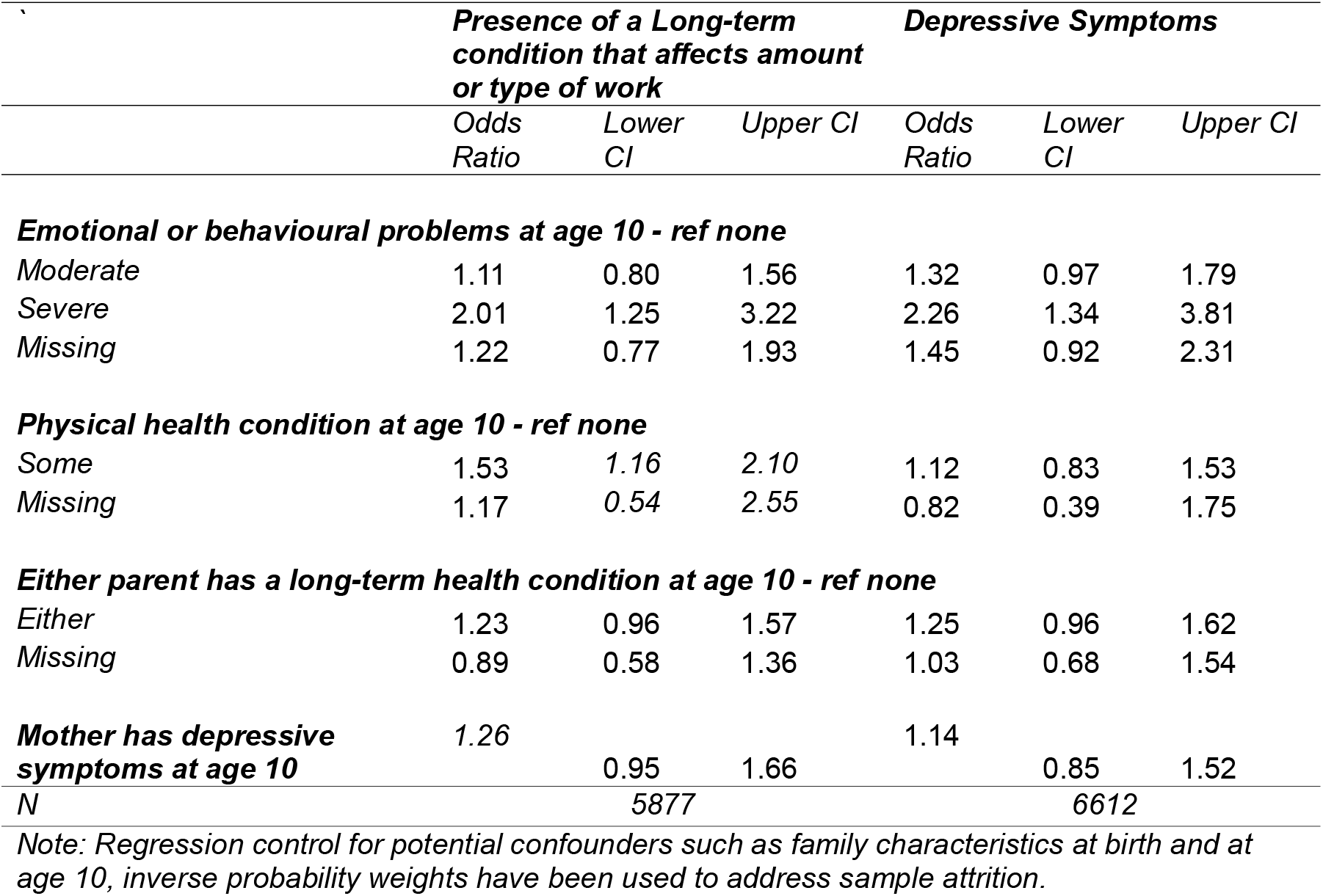
Odds ratios from logistic regressions of correlating adult health at age 51 and childhood health and family health at age 10.

## Discussion

Using a large, nationally representative longitudinal dataset, we provide evidence that poor physical and mental health in childhood is correlated with having lasting effects on individuals’ health into midlife - up to 40 years later. These findings underscore the importance of prioritising prevention and early intervention in childhood to promote healthier outcomes across the life course.

Although it is well established that childhood health influences outcomes in adulthood, our study adds to the existing literature by using the latest wave of the 1970 British Cohort Study (BCS), which now follows participants up to the age of 51. This age group is of growing policy importance: the UK has a comparatively low rate of labour market participation among older workers in the OECD, with just 65% of those aged 55–64 in work – only just above the OECD average of 63% (OECD, 2024). As the government seeks to raise employment and economic participation across the population, ensuring this group remains healthy and able to work is crucial. Our findings suggest that addressing health inequalities from early life may be part of may be part of a longer-term solution to economic activity across the life course.

A key strength of this study lies in its use of a large, population-based, prospective cohort with detailed information on childhood health, emotional and behavioural difficulties at age 10, and rich sociodemographic data. This enables us to control for a wide range of potential confounders and to provide robust estimates of the long-term relationship between early-life health and adult outcomes.

However, several limitations should be noted. First, the findings are specific to individuals born in Britain in 1970 and may not generalise to other birth cohorts or countries. Additionally, the latest wave of data collection occurred during the COVID-19 pandemic, which may have influenced health outcomes and participants’ responses.

As with all observational studies, we cannot rule out bias due to unmeasured confounding. Longitudinal studies are susceptible to attrition over time. To address this, we used inverse probability weighting and mean imputation with missing-indicator dummies. This approach assumes that data are missing at random-an assumption supported by the fact that most missing-data indicators were not statistically significant predictors of our outcomes.

Another limitation relates to the measurement of child behaviour. At age 10, behavioural and emotional problems were reported by the child’s mother, and these assessments may have been influenced by the mother’s own mental health. Although we include controls for maternal malaise to address this, potential interaction effects between child and maternal mental health may not be fully accounted for.

Despite these limitations, our findings provide evidence of the potential long-term consequences of poor childhood health—highlighting the case for investment in early intervention, particularly for children experiencing mental health challenges. Such action could yield substantial benefits not only for individual wellbeing but also for long-term economic and public health outcomes.

## Data Availability

The main survey data from BCS70 are available from the UK Data Service (University College London, 2025)

---

**Table A1:** Variables used in models adjusting for sample attrition.

| **Variable** | **Description** |
| --- | --- |
| *Country of birth* |  |
| England | Dummy = 1 if born in England |
| Wales | Dummy = 1 if born in Wales |
| Scotland | Dummy = 1 if born in Scotland |
| Northern Ireland | Dummy = 1 if born in Northern Ireland |
| Missing | Dummy = 1 if birth missing |
| *Voted in 1997 election* |  |
| Voted | Dummy = 1 if voted |
| Did not vote | Dummy = 1 if did not vote |
| Missing voted | Dummy = 1 if missing vote |
| *Parity* | *Number of older children in household at birth* |
| 0 | Dummy = 1 if number is zero |
| 1 | Dummy = 1 if number is one |
| 2 | Dummy = 1 if number is two |
| 3 | Dummy = 1 if number is three |
| 4 or more | Dummy = 1 if number is four or more |
| Missing | Dummy = 1 if number if missing |
| *Marital status at birth* |  |
| Married | Dummy = 1 if married |
| Not married | Dummy = 1 if not married |
| Missing | Dummy = 1 if missing |
| *Fathers level of education at birth* |  |
| Left before age of 15 | Dummy = 1 if below age of 15 |
| 15 to 18 | Dummy = 1 if above 15 but below 18 |
| 18+ | Dummy = 1 if above 18 |
| Missing level of education | Dummy = 1 if missing |
| *Contraception* |  |
| None | Dummy = 1 if none taken |
| Pill | Dummy = 1 if pill |
| Pill and or Other | Dummy = 1 if pill and or other method |
| Missing | Dummy = 1 if missing |
| *Pre-marital conception* |  |
| Yes | Dummy = 1 if out of wedlock |
| No | Dummy = 1 if not out of wedlock |
| Missing | Dummy = 1 if missing |
| *Socio-economic status of father* |  |
| I | Dummy = 1 if SC 1 |
| II | Dummy = 1 if SC 2 |
| III | Dummy = 1 if SC 3 |
| IV | Dummy = 1 if SC 4 |
| V | Dummy = 1 if SC 5 |
| VI | Dummy = 1 if SC 6 |
| Missing | Dummy = 1 if SC missing |
| *Number of house moves before age 5* | *Number of house moves before age of 5* |
| Missing number of house moves | Dummy = 1 if house moves missing |
| *Test Score* |  |
| Copy and Design score age 5 | Copy and design test score at age 5 |
| Missing Copy and Design score age 5 | Dummy = 1 if Copy and design test score at age 5 |
| BAS score age 10 | BAS score at age 10 |
| Missing BAS score age 10 | Dummy = 1 if Copy and design test score at age 5 |

**Table A2:**
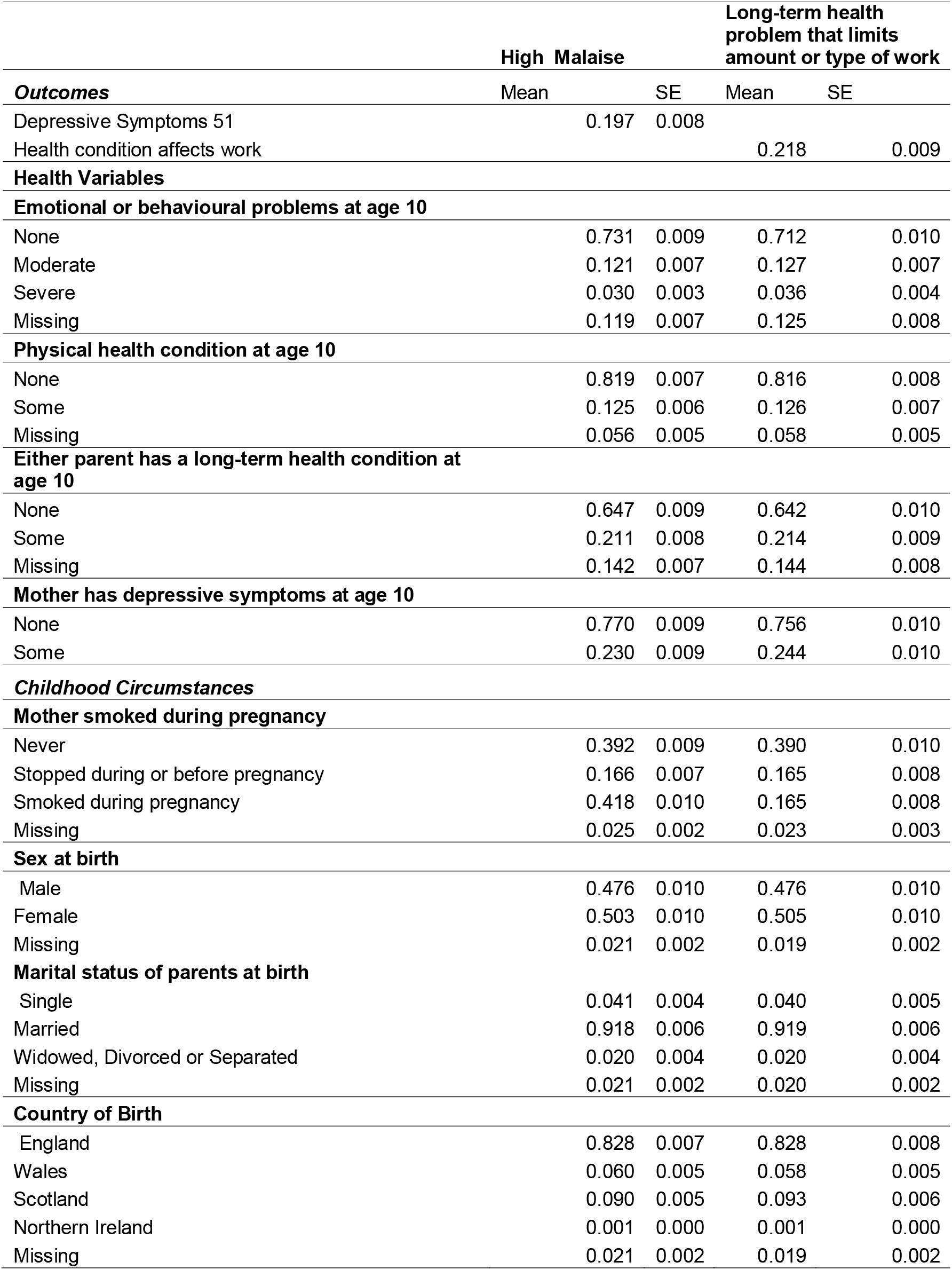
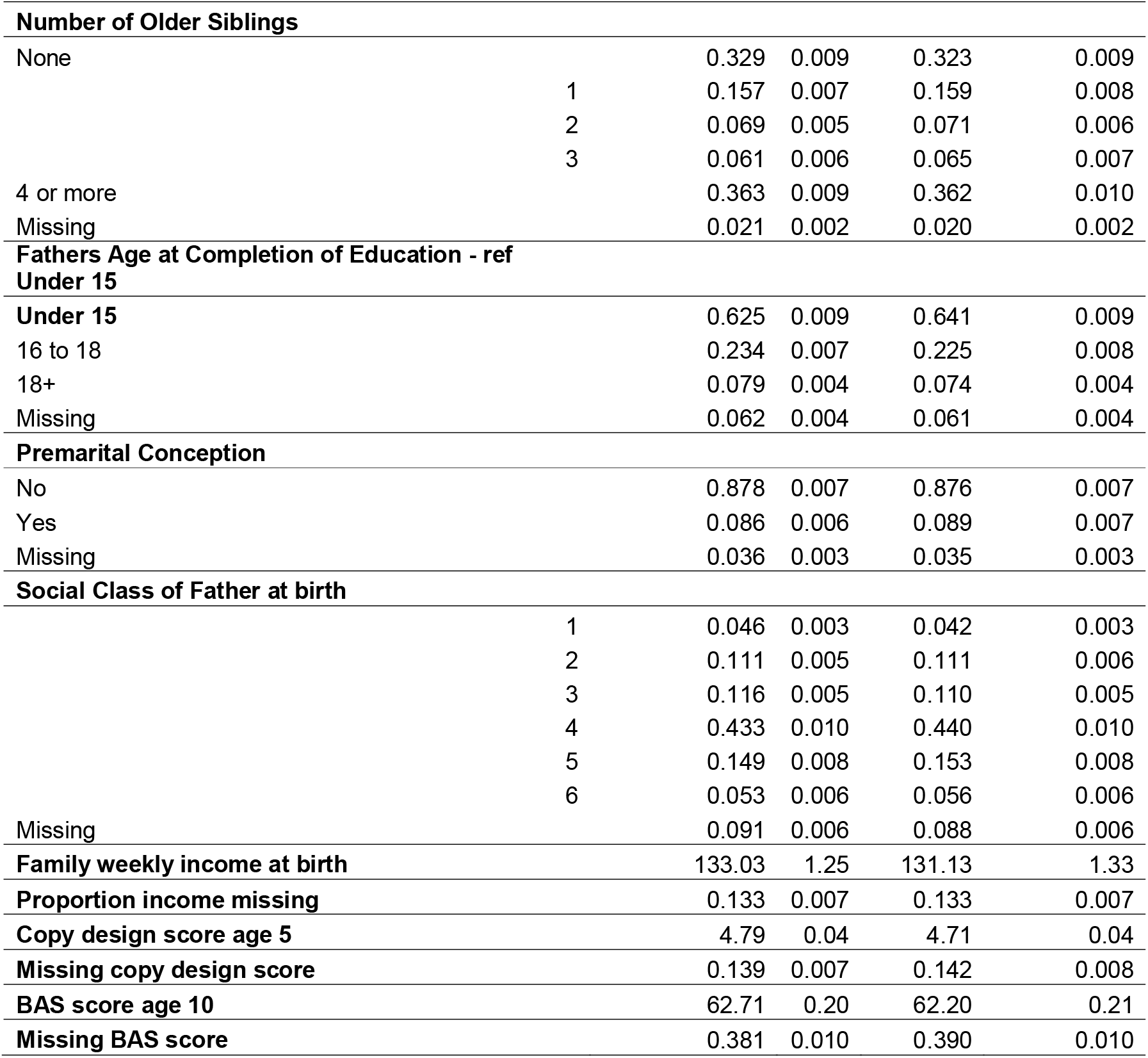
Variables used in models adjusting for sample attrition.

| **Outcomes** | **High Malaise** |  | **Long-term health problem that limits amount or type of work** |  |
| --- | --- | --- | --- | --- |
|  | Mean | SE | Mean | SE |
| Depressive Symptoms 51 | 0.197 | 0.008 |  |  |
| Health condition affects work |  |  | 0.218 | 0.009 |
| **Health Variables** |  |  |  |  |
| **Emotional or behavioural problems at age 10** |  |  |  |  |
| None | 0.731 | 0.009 | 0.712 | 0.010 |
| Moderate | 0.121 | 0.007 | 0.127 | 0.007 |
| Severe | 0.030 | 0.003 | 0.036 | 0.004 |
| Missing | 0.119 | 0.007 | 0.125 | 0.008 |
| **Physical health condition at age 10** |  |  |  |  |
| None | 0.819 | 0.007 | 0.816 | 0.008 |
| Some | 0.125 | 0.006 | 0.126 | 0.007 |
| Missing | 0.056 | 0.005 | 0.058 | 0.005 |
| **Either parent has a long-term health condition at age 10** |  |  |  |  |
| None | 0.647 | 0.009 | 0.642 | 0.010 |
| Some | 0.211 | 0.008 | 0.214 | 0.009 |
| Missing | 0.142 | 0.007 | 0.144 | 0.008 |
| **Mother has depressive symptoms at age 10** |  |  |  |  |
| None | 0.770 | 0.009 | 0.756 | 0.010 |
| Some | 0.230 | 0.009 | 0.244 | 0.010 |
| **Childhood Circumstances** |  |  |  |  |
| **Mother smoked during pregnancy** |  |  |  |  |
| Never | 0.392 | 0.009 | 0.390 | 0.010 |
| Stopped during or before pregnancy | 0.166 | 0.007 | 0.165 | 0.008 |
| Smoked during pregnancy | 0.418 | 0.010 | 0.165 | 0.008 |
| Missing | 0.025 | 0.002 | 0.023 | 0.003 |
| **Sex at birth** |  |  |  |  |
| Male | 0.476 | 0.010 | 0.476 | 0.010 |
| Female | 0.503 | 0.010 | 0.505 | 0.010 |
| Missing | 0.021 | 0.002 | 0.019 | 0.002 |
| **Marital status of parents at birth** |  |  |  |  |
| Single | 0.041 | 0.004 | 0.040 | 0.005 |
| Married | 0.918 | 0.006 | 0.919 | 0.006 |
| Widowed, Divorced or Separated | 0.020 | 0.004 | 0.020 | 0.004 |
| Missing | 0.021 | 0.002 | 0.020 | 0.002 |
| **Country of Birth** |  |  |  |  |
| England | 0.828 | 0.007 | 0.828 | 0.008 |
| Wales | 0.060 | 0.005 | 0.058 | 0.005 |
| Scotland | 0.090 | 0.005 | 0.093 | 0.006 |
| Northern Ireland | 0.001 | 0.000 | 0.001 | 0.000 |
| Missing | 0.021 | 0.002 | 0.019 | 0.002 |

|  |  |  |  |  |  |
| --- | --- | --- | --- | --- | --- |
| **Number of Older Siblings** |  |  |  |  |  |
| None |  | 0.329 | 0.009 | 0.323 | 0.009 |
|  | 1 | 0.157 | 0.007 | 0.159 | 0.008 |
|  | 2 | 0.069 | 0.005 | 0.071 | 0.006 |
|  | 3 | 0.061 | 0.006 | 0.065 | 0.007 |
| 4 or more |  | 0.363 | 0.009 | 0.362 | 0.010 |
| Missing |  | 0.021 | 0.002 | 0.020 | 0.002 |
| **Fathers Age at Completion of Education - ref Under 15** |  |  |  |  |  |
| **Under 15** |  | 0.625 | 0.009 | 0.641 | 0.009 |
| 16 to 18 |  | 0.234 | 0.007 | 0.225 | 0.008 |
| 18+ |  | 0.079 | 0.004 | 0.074 | 0.004 |
| Missing |  | 0.062 | 0.004 | 0.061 | 0.004 |
| **Premarital Conception** |  |  |  |  |  |
| No |  | 0.878 | 0.007 | 0.876 | 0.007 |
| Yes |  | 0.086 | 0.006 | 0.089 | 0.007 |
| Missing |  | 0.036 | 0.003 | 0.035 | 0.003 |
| **Social Class of Father at birth** |  |  |  |  |  |
|  | 1 | 0.046 | 0.003 | 0.042 | 0.003 |
|  | 2 | 0.111 | 0.005 | 0.111 | 0.006 |
|  | 3 | 0.116 | 0.005 | 0.110 | 0.005 |
|  | 4 | 0.433 | 0.010 | 0.440 | 0.010 |
|  | 5 | 0.149 | 0.008 | 0.153 | 0.008 |
|  | 6 | 0.053 | 0.006 | 0.056 | 0.006 |
| Missing |  | 0.091 | 0.006 | 0.088 | 0.006 |
| **Family weekly income at birth** |  | 133.03 | 1.25 | 131.13 | 1.33 |
| **Proportion income missing** |  | 0.133 | 0.007 | 0.133 | 0.007 |
| **Copy design score age 5** |  | 4.79 | 0.04 | 4.71 | 0.04 |
| **Missing copy design score** |  | 0.139 | 0.007 | 0.142 | 0.008 |
| **BAS score age 10** |  | 62.71 | 0.20 | 62.20 | 0.21 |
| **Missing BAS score** |  | 0.381 | 0.010 | 0.390 | 0.010 |

**Table A3:** Variables used in models adjusting for sample attrition.

|  | **Long-term condition that affects amount or type of work** |  |  | **Depressive Symptoms** |  |  |
| --- | --- | --- | --- | --- | --- | --- |
|  | Odds Ratio | Lower | Upper | Odds Ratio | Lower | Upper |
| **Emotional or behavioural problems at age 10 - ref none** |  |  |  |  |  |  |
| Moderate | 1.09 | 0.78 | 1.53 | 1.31 | 0.96 | 1.78 |
| Severe | 2.00 | 1.24 | 3.23 | 2.25 | 1.34 | 3.80 |
| Missing | 1.22 | 0.77 | 1.92 | 1.46 | 0.92 | 2.32 |
| **Physical health condition at age 10 - ref none** |  |  |  |  |  |  |
| At least one | 1.56 | 1.16 | 2.10 | 1.14 | 0.84 | 1.55 |
| Missing | 1.17 | 0.54 | 2.55 | 0.84 | 0.39 | 1.77 |
| **Either parent has a long-term health condition at age 10 - ref none** |  |  |  |  |  |  |
| Either | 1.24 | 0.97 | 1.58 | 1.26 | 0.97 | 1.63 |
| Missing | 0.87 | 0.57 | 1.33 | 1.01 | 0.67 | 1.51 |
| **Mother has depressive symptoms at age 10** | 1.23 | 0.93 | 1.63 | 1.12 | 0.84 | 1.50 |
| **Mother smoked during pregnancy - ref never** |  |  |  |  |  |  |
| Stopped during or before pregnancy | 1.54 | 1.13 | 2.09 | 1.12 | 0.83 | 1.50 |
| Smoked during pregnancy | 1.51 | 1.20 | 1.91 | 1.54 | 1.22 | 1.94 |
| Missing | 0.37 | 0.10 | 1.39 | 0.90 | 0.32 | 2.53 |
| **Sex at birth - ref Male** |  |  |  |  |  |  |
| Female | 1.55 | 1.24 | 1.93 | 2.02 | 1.61 | 2.53 |
| Missing | 0.18 | 0.01 | 4.30 | 0.42 | 0.01 | 12.66 |
| **Marital status of parents at birth - ref Single** |  |  |  |  |  |  |
| Married | 1.00 | 0.46 | 2.19 | 1.00 | 0.44 | 2.26 |
| Widowed, Divorced or Separated | 1.20 | 0.41 | 3.53 | 1.26 | 0.40 | 4.00 |
| Missing | 5.88 | 0.53 | 65.43 | 1.75 | 0.07 | 41.57 |
| **Country of Birth - ref England** |  |  |  |  |  |  |
| Wales | 0.88 | 0.57 | 1.36 | 1.16 | 0.78 | 1.71 |
| Scotland | 0.67 | 0.47 | 0.96 | 0.92 | 0.64 | 1.33 |
| Northern Ireland | 0.37 | 0.04 | 3.10 | 0.20 | 0.02 | 2.00 |
| Missing | 1.00 |  |  | 1.00 |  |  |
| **Log household family equivalised income at age 10** | 0.64 | 0.51 | 0.81 | 0.68 | 0.54 | 0.85 |
| Missing household income at age 10 | 0.87 | 0.59 | 1.29 | 0.99 | 0.67 | 1.46 |
| **Social Class of Father at birth - ref 1** |  |  |  |  |  |  |
| 2 | 1.12 | 0.69 | 1.82 | 0.88 | 0.57 | 1.37 |
| 3 | 1.08 | 0.66 | 1.75 | 0.87 | 0.56 | 1.36 |
| 4 | 1.12 | 0.71 | 1.76 | 0.85 | 0.56 | 1.30 |
| 5 | 1.21 | 0.72 | 2.03 | 0.94 | 0.57 | 1.53 |
| 6 | 1.33 | 0.70 | 2.55 | 1.01 | 0.54 | 1.91 |
| Missing | 2.15 | 1.12 | 4.12 | 1.01 | 0.53 | 1.96 |
| **Number of Older Siblings - ref none** |  |  |  |  |  |  |
| 1 | 0.94 | 0.71 | 1.25 | 1.03 | 0.78 | 1.35 |
| 2 | 0.96 | 0.68 | 1.36 | 1.06 | 0.75 | 1.50 |
| 3 | 1.12 | 0.73 | 1.71 | 0.95 | 0.62 | 1.45 |
| 4 or more | 1.09 | 0.65 | 1.83 | 0.79 | 0.44 | 1.40 |
| Missing | 1.12 | 0.15 | 8.46 | 1.35 | 0.27 | 6.76 |
| **Fathers Age at Completion of Education - ref Under 15** |  |  |  |  |  |  |
| 16 to 18 | 0.71 | 0.55 | 0.92 | 0.87 | 0.67 | 1.15 |
| 18+ | 0.64 | 0.43 | 0.94 | 0.74 | 0.52 | 1.05 |
| Missing | 1.07 | 0.62 | 1.86 | 0.95 | 0.54 | 1.67 |
| **Premarital Conception - ref No** |  |  |  |  |  |  |
| Yes | 1.09 | 0.70 | 1.70 | 1.16 | 0.76 | 1.78 |
| Missing | 1.12 | 0.58 | 2.18 | 1.58 | 0.87 | 2.84 |
| **Copy design score at 5** | 0.93 | 0.87 | 0.99 | 0.98 | 0.92 | 1.04 |
| **Missing design score 5** | 1.33 | 0.99 | 1.79 | 1.32 | 0.98 | 1.79 |
| **BAS score 10** | 1.00 | 0.98 | 1.01 | 0.99 | 0.97 | 1.00 |
| **Missing BAS Score** | 0.98 | 0.78 | 1.22 | 1.31 | 1.05 | 1.63 |
| Constant | 1.39 | 0.28 | 6.94 | 1.21 | 0.23 | 6.28 |
| N |  | 5877 |  | 6612 |  |  |

**Table A4:**
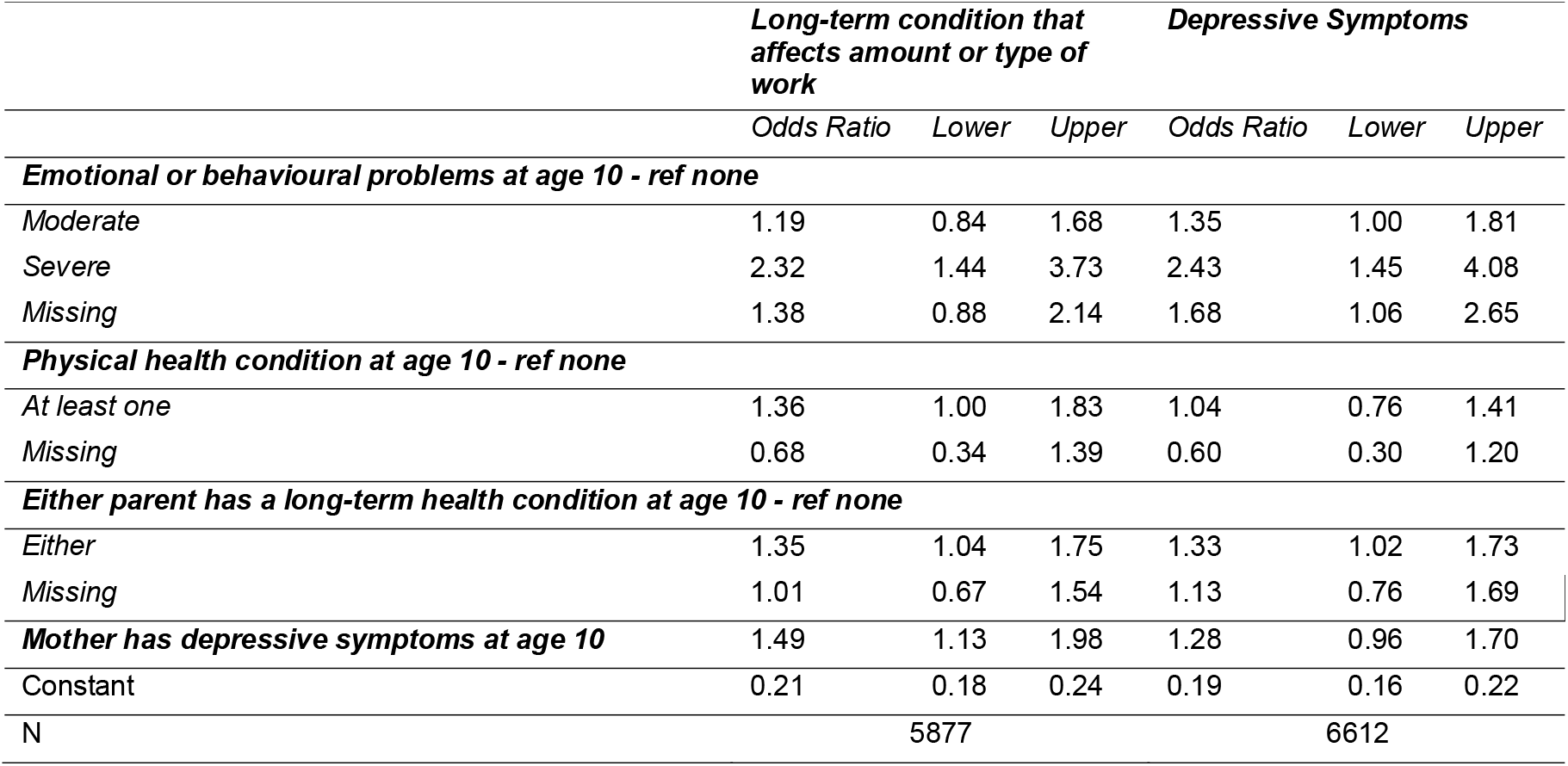
Variables used in models adjusting for sample attrition.

